# Demonstrate, observe, assist perform (DOAP) versus structured educational video (SEV) in imparting standard skill in male urinary bladder catheterisation

**DOI:** 10.1101/2021.03.06.21250996

**Authors:** Santosh Balakrishnan, Lijo Paul, Minu N Rajan, Sherin A Arthungal

## Abstract

**Introduction:** Conventional teaching and learning methods have been seen to fail to assure achievement of competencies in male bladder catherization in the Indian Medical Graduate (IMG) with wide variation noted in competencies. This could be remedied by introduction of Structured training methods.

**Aims and Objective:** The study aimed to investigate DOAP method of training in a Skill Lab against training through a Structured Educational Video (SEV) with the objective of comparing their efficacy in training Final MBBS Students in the psychomotor skill of performing male bladder catheterisation.

**Material & Methods:** Final MBBS students fulfilling selection criteria were randomly allocated into two comparable groups. One group underwent Skill lab training using DOAP method while the other group underwent training using a SEV by the same instructor. CRRI interns, regularly performing MBC at work by virtue of conventional training, with 6-8 months experience formed a control group. All participants underwent assessment of skill in MBC by skill lab OSCE evaluation, by assessors who were blinded to the participant’s method of training. Data was recorded and analysed using standard statistical software. Trial evaluation from the trial groups was obtained using Survey monkey tool.

**Observation:** There was no statistically significant difference in the ability of DOAP group or SEV group in being able to safely perform MBC though a higher level of confidence was expressed with their training by DOAP group. Both trial groups statistically outperformed the control group.

**Conclusion:** Structured training assures competence. Video-assisted Training produces comparable results though DOAP method is preferred by students. A combination of the techniques may facilitate optimal training.

## Introduction

Wide variation is observed in skills in performing common bedside clinical procedure of male bladder catheterisation among final phase MBBS students trained through conventional training and assessment [1,2]. Conventional teaching and learning methods may not allow assurance of achievement of competencies in the Indian Medical Graduate IMG. Introduction of structured training methods could offer a reliable method of facilitation of learning and assured skill acquisition.

## Aim

The team aimed to investigate the suitability of Demonstrate, observe, assist perform (DOAP) method of training in a Skill Lab simulator compared with demonstration using a Structured Educational Video (SEV) in imparting psychomotor skill training in a bedside procedure to medical students.

## Objective

The objective of this trial was to evaluate the efficacy of training by DOAP in comparison to training through a SEV in acquiring skill in performing male bladder catheterisation among Final phase MBBS Students

## Material & Methods

The study was performed as a mandated research project as part of the 1-year Advanced course in Medical Education administered by the National Medical Council through its nodal centre in Kottayam, Kerala. The total duration of the trial was therefore restricted by course regulation to 6 months between conceptualisation & presentation of results as a poster to the course peer group & faculty.

The study was designed as a randomised comparative study conducted in the investigator’s institution. A SEV was created mirroring the steps and instructions conveyed by the DOAP training session. This SEV was approved through peer review by four consultant surgeons, one of whom was a consultant Urologist. The DOAP session & OSCE assessment of all groups was conducted in a simulation lab with procedure demonstrated and assessed using a standard male catheterisation trainer model. The group being trained through a SEV were trained by being shown the video twice in the seminar room in the presence of the trainer who led the training session and also reinforced the steps by verbal repetition between two successive viewings of the SEV.

The larger population of the study are Final phase MBBS Students of Ernakulam district in Kerala, India.

### Sampling method

All Final phase MBBS students fulfilling selection criteria were randomly assigned a trial number by blinded chit pick-up for anonymity with a securely held key for verification if necessary. They were divided into two equal groups based on random selection and grouping again by blinded chit pick-ups. Compulsory Residential Rotating Internship (CRRI) interns were recruited through their random allocation to the department as per their rotation, to serve as a control group. The interns posted to the department for the duration of the study would have completed between six to eight months of CRRI with regular opportunity at work to perform MBC. They represent a comparable group who have just completed final phase MBBS and have learnt the procedure through the conventional apprenticeship method.

### Inclusion Criteria

1. Medical Students from Final Phase who have not had skill training in male bladder catheterisation who consent to participate
2. CRRI interns posted to the department of surgery subject to their consent.

### Exclusion Criteria

1. Medical students who have had structured skill training through any means in bladder catheterisation.
2. CRRI interns who had any structured training in bladder catheterisation other than traditional apprenticeship.
3. CRRI interns from the senior additional batch.
4. Subjects who have Allergy to latex.

### Sample size

All final phase MBBS Students who fulfil inclusion criteria were recruited. The number of participants was limited by the maximum number of students fulfilling criteria in the institution over the permitted study period.

### Intervention

Institutional review board and Ethical Committee clearance and written informed consent from all participants was taken. The DOAP group underwent structured training using the DOAP method in batches of not more than 6 subjects with demonstration and observation as a group and individual opportunity to assist and perform. The group being trained through a SEV was trained in batches of 15, as limited by the size of the seminar room available for screening the video. They were shown the video two times in the presence of the trainer who verbally reinforced the steps between viewings, to match the visual and auditory exposure through DOAP. A group of 35 CRRI interns posted in our department in numbers to reasonably match the test groups, act as controls trained by conventional apprenticeship method of training to verify the validity and impact of either of the study methods of training over current practice as recommended by members of the institutional research committee. Participants in all groups were individually assessed and scored using an existing validated OSCE assessment form [3] for their skill in male catheterisation by assessors blinded to the intervention in question. The OSCE form assessed completion of 26 vital steps of the procedure. Eight vital steps were allocated two marks while all others had one mark making up a total score of 34. Following assessments, an online programme satisfaction evaluation questionnaire was administered to each participant. Debrief to subjects was provided only after collection of evaluation questionnaire. In the interest of fairness, a crossover training was provided by sending the video to all in the DOAP group and DOAP training was offered to all in the SEV group after the assessments and online evaluation was complete in order to avoid bias in response.

### Data Collection Method and Analysis

OSCE valuation scores in whole numbers for all groups were compiled in excel sheets. SPSS software was used for data analysis. The institutional bio-statistician aided the analysis in view of the authors’ limited skill with the software tool. P<0.05 was considered as statistically significant. OSCE scores were summarised using appropriate tests of significance using these tools. Evaluation response received from both groups using Survey Monkey online survey tool, was analysed for difference in levels of agreement on various elements of the training experience with their allocated training method after they completed the OSCE assessment. Data was analysed using MS Excel.

## Results

All trial intervention groups; DOAP group (n=39), SEV group (n=39) and the control group participants (n=35) successfully completed catheterisation in the Simulator model in the skills lab. OSCE scores were awarded by assessors blinded to the interventions. Weight given to each of the 26 elements in the OSCE assessment as well as total scores out of 34 were recorded on proformas and compiled on MS Excel. The scores were found to follow normal distribution (Kolmogorov-Smirnov test) in control, DOAP and SEV groups. (**Fig 1**) Means and standard deviations were calculated. Initial One-way Anova was performed using SPSS software to assess significant difference in the mean scores for each of the three groups. Fischer’s exact test showed (F value=65.53; P<0.001) significant difference between the three groups.

**Fig 1:**
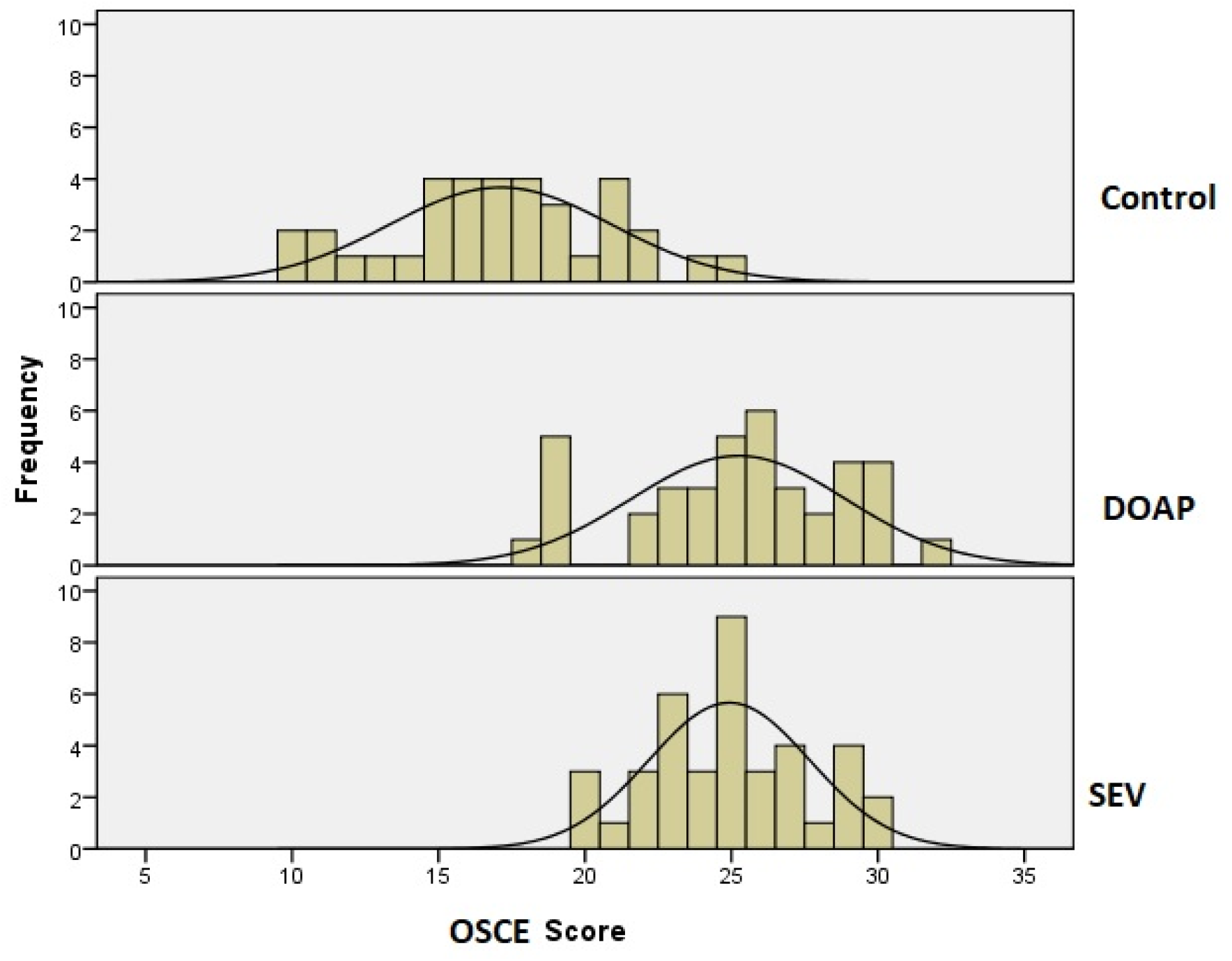
Histogram of OSCE results: Normal Distribution.

In keeping with the objective of the study a Post Hoc Test (Tukey-HSD) performed to compare the DOAP and SEV groups was done. There was no significant difference between the mean scores of the two groups (P=0.92). There was significant difference however between SEV and DOAP groups individually against the control group (**Table 1**).

**Table 1:** Anova Comparison of groups.

| OSCE Score Results |  |  | Three-way Anova |  | One Way Anova with Tukey HSD Post Hoc Inter Group Comparison |  |  |  |
| --- | --- | --- | --- | --- | --- | --- | --- | --- |
| Group | n= Numbers | Mean OSCE Score | F Value | P- Value. | Comparison of Groups | Mean Difference | P Value | Significant Favouring |
| Control | 35 | 17.11 | 65.528 | <0.001 | DOAP v/s SEV | 0.308 | **0.917** | Neither |
| DOAP | 39 | 25.23 |  |  | Control v/s DOAP | -8.116 | **<0.001** | DOAP |
| SEV | 39 | 24.92 |  |  | Control v/s SEV | -7.809 | **<0.001** | SEV |

Assessment of the feedback response on SurveyMonkey showed a statistically significant difference in the level of agreement with all elements of training surveyed favouring the DOAP group (**Table 2**). They reported higher levels of satisfaction with the method of training, felt it reinforced elements of risk, safety and caution involved in performing the procedure and also strongly recommended it as a modality of training for trainees at their level. The video group reported lower levels of satisfaction with the confidence instilled by the training process in performing the procedure and were less convinced of the suitability of video assisted training as a method of psychomotor training for trainees at their level of expertise.

**Table 2:** Analysis of Study subject’s Trial experience feedback (Surveymonkey®).

| Questions presented | Groups | Numbers in Strong agreement | Percentage in Strong Agreement | P- Value: Significant if P< 0.05 |
| --- | --- | --- | --- | --- |
| 1. Captivating & Promotes interest | DOAP | 25 | 73.5 | 0.002 |
|  | SEV | 12 | 34.3 |  |
| 2. Clear explanation of Procedure | DOAP | 25 | 73.5 | 0.007 |
|  | SEV | 14 | 40 |  |
| 3. Elements of risk and safety conveyed | DOAP | 18 | 52.9 | 0.045 |
|  | SEV | 9 | 25.7 |  |
| 4. Confidence afforded to perform | DOAP | 17 | 50 | <0.001 |
|  | SEV | 4 | 11.4 |  |
| 5. Suitability of teaching method | DOAP | 26 | 76.5 | <0.001 |
|  | SEV | 5 | 14.3 |  |
| 6. Whether they would recommend this method | DOAP | 25 | 73.5 | <0.001 |
|  | SEV | 3 | 8.6 |  |
| 7. Overall satisfaction with teaching method | DOAP | 26 | 76.5 | <0.001 |
|  | SEV | 7 | 20 |  |
| Degree of affirmative agreement with the teaching method was significantly higher in the DOAP Group with a P value of <0.05 in every element. |  |  |  |  |

In a sub-analysis 8 critical elements of the procedure assessed in the OSCE (Questions 3, 6,10,14, 17, 18, 22, 26) were compiled (**Table 3**). Failure to perform each critical element of the procedure was compared between DOAP and SEV groups. It was observed, that contrary to the respondent’s feedback, the SEV group actually outperformed the DOAP group in the elements of the OSCE (Q3,10,17,18) that pertained to safety and risk mitigation in the procedure. They performed poorly in the steps (Q6,14, 22 & 26) that pertained to pre-procedure hand wash, adequate use of lubricant anaesthetic gel, post procedure replacement of prepuce to normal position and documentation in the notes. The difference failed to achieve statistical significance in any except Q22, pertaining to repositioning of prepuce post procedure, which favoured of the DOAP group.

**Table 3.** Subgroup analysis: Scores in 8 critical steps of procedure.

| Critical Steps assessed by OSCE | Fail | DOAP | SEV | P- Value Significant if P<0.05 |
| --- | --- | --- | --- | --- |
|  | Pass |  |  |  |
| Q3: Taking Consent | Fail | 7 | 4 | 0.520 |
|  | Pass | 32 | 35 |  |
| Q6: Hand washing pre-procedure | Fail | 1 | 5 | 0.200 |
|  | Pass | 38 | 34 |  |
| Q10: Clean by no-touch technique | Fail | 6 | 3 | 0.480 |
|  | Pass | 33 | 36 |  |
| Q14: Insertion of adequate anaesthetic gel | Fail | 1 | 3 | 0.620 |
|  | Pass | 38 | 36 |  |
| Q17: No-touch catheter insertion | Fail | 2 | 0 | 0.490 |
|  | Pass | 37 | 39 |  |
| Q18: Confirming Urine flow | Fail | 11 | 5 | 0.092 |
|  | Pass | 28 | 34 |  |
| Q22: Retraction of prepuce post procedure* | Fail | 17 | 28 | 0.012 |
|  | Pass | 22 | 11 |  |
| Q26: Documentation | Fail | 6 | 10 | 0.260 |
|  | Pass | 33 | 29 |  |
| * Bias Correction<br>Q22 discounted because MBC simulator had a circumcised penile shaft & hence could not be reliably assessed on OSCE. |  |  |  |  |

Despite the significant lower level of confidence expressed by the SEV group, there was no statistically significant difference in their ability to perform the procedure safely vis-à-vis the DOAP group as seen by their OSCE score assessment.

Each of the intervention groups outperformed the Control group with the difference reaching statistical significance.

### Inherent Bias & Redressal

1. Researcher’s interest in Structured competency-based training:
  - Excluded by recruiting two trial blinded assessors for each group assessment.
2. Repetition bias for DOAP group:
  - Minimised by limiting feedback to after completing OSCE assessment and submitting evaluation questionnaire.
  - SEV group shown video two times in presence of trainer to match visual impact with opportunity to discuss steps with trainer to match opportunity afforded to DOAP group.

## Discussion

The training of a medical graduate requires development of almost every domain of learning. Most core competencies require the application of more than one skill domain. The move towards competency based medical education (CBME) globally had highlighted this need and also the challenges inherent to ensuring achievement of these competencies [1]. Wide variability in procedural skills have been observed among junior doctors even after adoption of CBME raising questions about their readiness to undertake roles expected of them [2].

Simulation based training provides an evidence-based solution to imparting skills to the modern trainee in the current practise backdrop of reduced availability of trainee patient contact time as a result of reduced inpatient stay for most treatments and reduced tolerance for training related morbidity [4]. Skill labs allow repeated training and assessment in a safe environment till achievement of skills in objectively demonstrated before advancing to the clinical settings. Peyton’s 4 step technique (DOAP) is an effective method of psychomotor skill training. This technique has been shown to be significantly effective in procedures involving multiple sequential steps [5,6].

Many studies have confirmed the positive significant benefit of adding multimedia-based training to traditional text-based training [7]. Students have also shown grater satisfaction with the use of video assisted learning in comparison with traditional methods of skill training through demonstrations [8]. No significant difference was noted in procedural skill when video-based teaching was compared to live demonstrations for training in orthodontal procedures [8]. Students report personal comfort, availability of media for review and clarification and scope for personal visualisation and reflection as possible reasons for this preference [8,9].

The present study showed that, though the subjects trained through DOAP reported greater satisfaction and confidence with their training as compared to those trained through the use of a SEV, there was no significant difference in their OSCE performance scores in the skill lab. There was no statistical difference even in the analysis of 8 critical steps selected from the OSCE questionnaire apart from the step requiring the subject to declare the need to protract the prepuce post catheterisation in uncircumcised individuals where the difference reached statistical significance favouring the DOAP group. The DOAP group also had a numerical advantage over the SEV group in performing documentation of the procedure, though the difference didn’t reach statistical significance.

This marginal advantage could be accounted to the simulator model featuring a circumcised penis and the need to document the procedure being conveyed only as a closing statement by the narrator in the SEV rather than a scene of this being done.

The lack of visual impact of both these steps could have resulted in failure on the part of the subject to register the verbal suggestion impressing the need to protract the prepuce post catheterisation in uncircumcised males and ensure documentation of the procedure in the case notes. Including visuals of these steps being performed in the training video could potentially level the field.

Video based training thus appears to be as effective a tool when compared to the DOAP method for training in simple bedside clinical procedures.

There is evidence that structured training improves performance in healthcare professionals in procedural, communication and clinical examination skills over learning by apprenticeship and informal workplace-based learning [10,11].

The present study findings also show a clear statistical advantage in the OSCE scores achieved by subjects trained through either method of structured training (DOAP or SEV) over the subjects in the control group who were trained by the conventional way through apprenticeship and unstructured work-based supervised training.

## Conclusion

Structured training appears to be a key element in implementing CBME to ensure development of competencies. Personalised training through DOAP in small groups appears to be the ideal method of imparting training in common bedside clinical procedures but needs more time and faculty hours. Training using Structured educational videos with the presence and reinforcement by a trainer allows training of larger groups and appears to be equally effective. A combination of the techniques could facilitate training through optimal use of faculty & simulation lab resource without compromising learning.

## Data Availability

The author holds the data pertaining to the results of the study.

## Abbreviations

SEV: Structured Educational Video
DOAP: Demonstrate, Observe, Assist, Perform
CRRI: Compulsory Residential Rotating Internship
MBC: Male bladder catheterisation
IMG: Indian Medical Graduate

## Acknowledgment

1. Administration of MOSC Medical College; Dean, Dr KK Divaker. Medical Superintendent, Dr Sojan Ipe for permitting use of premises & skill lab for the study.
2. Faculty of the National Medical Council nodal centre, Kottayam & my peers from the Advanced course of Medical Education (ACME Batch 2019B)
3. My Colleagues: Dr Vergis, Dr Vijy, Dr Satish, Dr Shal & Dr Koruth, for their support & peer review of the SEV
4. Institutional Bio-Statistician: Mr Kalesh M Karun for his support with statistical analysis.
5. Our Interns: Batch of 2014 Our Students: Batch of 2015; Skill Lab nurse: Mr. Yaldho, Department Clerk: Ms Sini. Sister In-charge OT: Ms Anuja

## Notes

### Competing Interest Statement

The authors have declared no competing interest.

### Clinical Trial

The study was performed as a mandated research project as part of the 1-year Advanced course in Medical Education administered by the National Medical Council through its nodal centre in Kottayam, Kerala, India. Study was registered and approved by Institutional review board & Institutional Ethics committee: IRB Approval Date: 29/11/2019 IEC Protocol Number: MOSC/IEC/398/2019. My project was a simulation based educational research. Though it involved human volunteers, there was only an educational intervention to facilitate the learning of a bed side skill. No medications or physical interventions were involved. The outcome measure was based on ability to demonstrate the skill learnt in a simulation lab model and hence did not involve any direct impact on humans or patients. The training or absence of the same would have no impact on the formal academic assessment or progress of the volunteers outside of the trial environment. Fairness to the volunteers was ensured by offering each group the opportunity to experience the other educational intervention after assessment and feedback was complete. We believed therefore that the education research as described did not qualify as a clinical trial and hence registration as a clinical trial was not felt mandatory. The IRB & IEC permitted me to proceed with my work. I was under the impression then, that registration of such research though possible was not considered mandatory. Clinical Trials Registry of India unfortunately does not allow retrospective registrations.

### Funding Statement

The team did not receive any monetary funding. Acknowledgement to Mr Kalesh Karun, Biostatician, MOSC Medical College Hospital for help with Statistical analysis. Acknowledgement to administration of MOSC Medical College, for permitting use of Skill Lab and premises. Acknowledgement to my fellow learners of the Advanced Course in Medical Education: ACME Batch 2019B & the faculty of the National Medical Council Nodal center, Kottayam, Kerala for guidance & peer support. Acknowledgement to Dean: Dr KK Divaker. Medical Superintendent: Dr Sojan Ipe for institutional support. My surgical Colleagues: Dr Vergis, Dr Vigy, Dr Satish, Dr Shal & Dr Koruth for peer reviewing the Strutured video & support for the study; IRB in-charge: Dr Sara, IEC in-charge: Dr Sandhya Kurup for guidance. Skill Lab nurse: Br. Yaldho; Our Interns: Batch of 2014 Our Students: Batch of 2015 for agreeing to participate in the study. Department Clerk: Ms Sini for admin support. Sister In-charge of Operation theatre: Ms Anuja.

### Author Declarations

IRB Approval Date: 29/11/2019 IEC Protocol Number: MOSC/IEC/398/2019 Institutional Review Board & Ethics Committee. MOSC Medical College Hospital. Ernakulam. Kerala. India. 682311

### Summary of Updates

Minor modification to text of conclusion to mirror the words in the abstract without changing the context or findings. Tables 1 & 2 amalgamated to present the findings clearly and allowing comparison by readers

